# Chrono-optimization of influenza vaccine administration: A systematic review and meta-analysis

**DOI:** 10.1101/2024.02.15.24302880

**Authors:** Koen Vink, Jeroen Kusters, Jacco Wallinga

## Abstract

**Background:** There is growing evidence that the strength of vaccine responses depends on the time of day of vaccine administration. This systematic review provides an overview of the literature regarding the effect of the timing of influenza vaccination on the vaccine response. To estimate the extent of this effect, we conducted a meta-analysis of randomized controlled trials (RCTs) in which antigen-specific antibody titers were monitored following either morning or afternoon administration of the influenza vaccine.

**Methods and results:** A systematic literature search identified five relevant studies that reported antigen-specific titers against multiple influenza vaccine strains after both morning and afternoon vaccination. Four of the five studies reported higher antibody titers for at least one vaccine strain following morning vaccination. Two RCTs were included in the meta-analysis, each of which reported the response to three vaccine strains, resulting in a total of six responses. The meta-analysis revealed that morning vaccination elicited a stronger antibody response than afternoon vaccination, with a pooled standardized mean difference (SMD) of 0.24 (95% CI=0.01–0.47). The between-study heterogeneity (I^2^=66%) was mainly due to the significantly 01greater effect of morning vaccination among adults aged 65 years or older than among adults aged 60 years or younger (SMD=0.32, 95% CI=0.21–0.43 versus SMD=0.00, 95% CI=−0.16–0.16, respectively).

**Conclusion:** Influenza vaccinations administered in the morning induced a stronger antibody response in adults aged 65 years or older, who represent a major target group for influenza vaccination programs. Therefore, chrono-optimization of influenza vaccination could offer a safe and simple strategy for enhancing vaccine effectiveness. The paucity of relevant studies suggests that accounting for the time of vaccine administration in future vaccination trials could provide valuable insights into the potential benefits of chrono-optimization strategies.

## Introduction

Improving the level of vaccine-induced protection against frequent infections associated with a risk of severe disease is essential for minimizing the burden of such infectious diseases. Conventional approaches to improve the immunogenicity of vaccines, such as optimizing antigen presentation or incorporating new adjuvants, are limited by time-consuming safety testing and increased risks of adverse events [1]. There is growing evidence suggesting that the strength of vaccine responses depends on the time of day at which vaccines are administered [2, 3]. Therefore, optimizing the time of vaccine administration might offer a safe and simple strategy for enhancing vaccine effectiveness.

The effect of timing on the vaccine response can be attributed to oscillations in various components of our immune system throughout the day. Circadian rhythms have been described in cytokine responses [4, 5], circulating leukocyte counts [4, 6], the activity of innate immune cells [5], sensitivity to pathogen-associated molecular patterns [5, 6], and the expression of genes involved in the functioning of adaptive immune cells [5, 7]. These oscillations are driven by cell-intrinsic circadian clocks, which are composed of so-called *clock* proteins that regulate and maintain 24-hour cycles in the expression of genes involved in the cellular function by coordinating transcriptional and translational feedback loops [8]. These circadian rhythms are believed to play important roles in causing variations in vaccine responses depending on the time of day of administration [2, 9].

Studies that investigated the effect of vaccination timing for various infectious diseases have reported varying results. Some studies have demonstrated that morning vaccination against influenza [10–13], hepatitis A [10], SARS-CoV-2 [14] and tuberculosis [15] induces a more robust antibody response than afternoon or evening vaccination. In contrast, other studies have reported stronger immune responses following vaccination later in the day for SARS-CoV-2 [16], or reported no differences between vaccinations administered at different timepoints for influenza [17], SARS-CoV-2 [18] and hepatitis B [19]. Thus, the optimal time of vaccination as well as the potential impact on the level of protection may depend on the vaccine.

This study focusses on the effect of influenza vaccination timing on the vaccine response. There are several reasons for focusing specifically on influenza. Firstly, influenza imposes a high disease burden, with an estimated 650,000 deaths annually worldwide [20], and an enormous economic burden due to costs related to the expenditure of medical resources and absenteeism from work [21, 22]. Secondly, there is room for improving the moderate effectiveness of influenza vaccines, especially among older adults who experience the highest rates of influenza-associated hospitalization and mortality [23, 24]. Finally, the majority of the existing evidence from randomized controlled trials (RCTs) regarding optimal vaccination timing pertains specifically to influenza vaccines. Since influenza vaccines are multivalent, there is also more data on the vaccine response compared to monovalent vaccines.

Here, we provide a comprehensive overview of the current evidence from observational studies and RCTs comparing influenza antibody titers following morning and afternoon vaccine administration. To estimate the magnitude of this time-of-day effect, we conducted a meta-analysis of available RCTs that compared strain-specific antibody titers one month after either morning or afternoon administration of the influenza vaccine.

## Methods

This study followed the Preferred Reporting Items for Systematic Reviews and Meta-Analyses (PRISMA) guidelines (Table S1) [25]. The review was not registered in PROSPERO.

### Literature search and study selection

To identify eligible studies for this review, a systematic literature search was conducted in PubMed, Embase, MEDLINE, Preprints, and the Cochrane Central Register of Controlled Trials (CENTRAL). A search string was created that contained key terms such as “influenza vaccination”, “time of day”, and “vaccination timing”; see S2. The databases were searched up to the 15^th^ of August 2023, without applying any filters. Studies that met the following criteria were included in the review: participants were free of current infections or immunodeficiencies, antigen-specific antibody titers or T-cell responses were measured following influenza vaccination and compared between participants vaccinated at different timepoints throughout the day. Subsequently, these studies were only included in the meta-analysis if the time of vaccine administration was randomized and if antigen-specific antibody titers were measured at least one month post vaccination. Two researchers independently conducted the study selection process. After discarding duplicates, all identified records were screened for eligibility based on the title and abstract. This was followed by a full text review to assess whether the remaining records met the aforementioned eligibility criteria for both the review and meta-analysis. If no consensus was reached for the study selection, a third researcher was consulted.

### Risk of bias assessment

The risk of bias of the included studies was critically appraised by two assessors using Cochrane’s Risk of Bias tool 2 (RoB2) for (cluster-)RCTs [26], and the Risk Of Bias In Nonrandomized Studies of Interventions (ROBINS-I) tool for observational studies [27]. A final consensus judgement was reached for each study by considering the evaluations of both assessors, and if necessary a third assessor was consulted. Publication bias was assessed by checking clinical trial registers for ongoing or unpublished studies.

### Data collection

The study type, location, number of subjects and their characteristics, vaccination type, vaccination time, and study outcomes were extracted from the included studies. The data required for the meta-analysis were collected from the published (supplementary) materials and by contacting the authors. These data included the mean and standard deviation (SD) of antigen-specific antibody titers measured one month after vaccination for the different influenza vaccine strains (A/H1N1, A/H3N2, and type B influenza). Reported titers were log-transformed to standardize them onto a common logarithmic scale. Henceforth, we will refer to these log-antibody titers as simply “antibody titers”.

### Outcomes

The primary outcome of the meta-analysis was the standardized mean difference (SMD) in antibody titers one month post vaccination between morning and afternoon vaccine administrations. The secondary outcomes were the effect of age and sex on the effect of vaccination timing on the antibody response. The group sizes and the means and SDs of the antibody titers were used to calculate the SMDs in titer levels between morning and afternoon vaccination, and the corresponding variances and standard errors per study using the “*metafor*” package [28] in the software environment for statistical computing: R, version 4.3.0 (R Core Team, Vienna, Austria).

### Statistical analysis

A three-level random-effects model was used to obtain a pooled effect estimate with confidence intervals for the difference in the antibody response between morning and afternoon vaccination. This model corrected for the correlation between the multiple effect sizes within each study. A forest plot was created using the model output to visualize the results of the meta-analysis. Heterogeneity between the selected studies was assessed by inspecting the forest plot and using the *tau^2^*, Cochran’s *Q* and *I^2^* statistics. Subsequently, subgroup analyses were conducted based on sex, age group, and vaccine strain. All analyses were performed in R (version 4.3.0) with the “*metafor*” [28] package.

## Results

Of the 217 records identified in the literature search, five met the eligibility criteria for inclusion in the systematic review (Figure 1) [10–13, 17, 29]. In every study, the participants were vaccinated with standard-practice trivalent (or quadrivalent) inactivated influenza vaccines in either the morning or afternoon. The characteristics and findings of the included studies are summarized in Table 1. Two of these five studies were RCTs [11–13], and were therefore also included in the meta-analysis (Figure 1).

**Figure 1:**
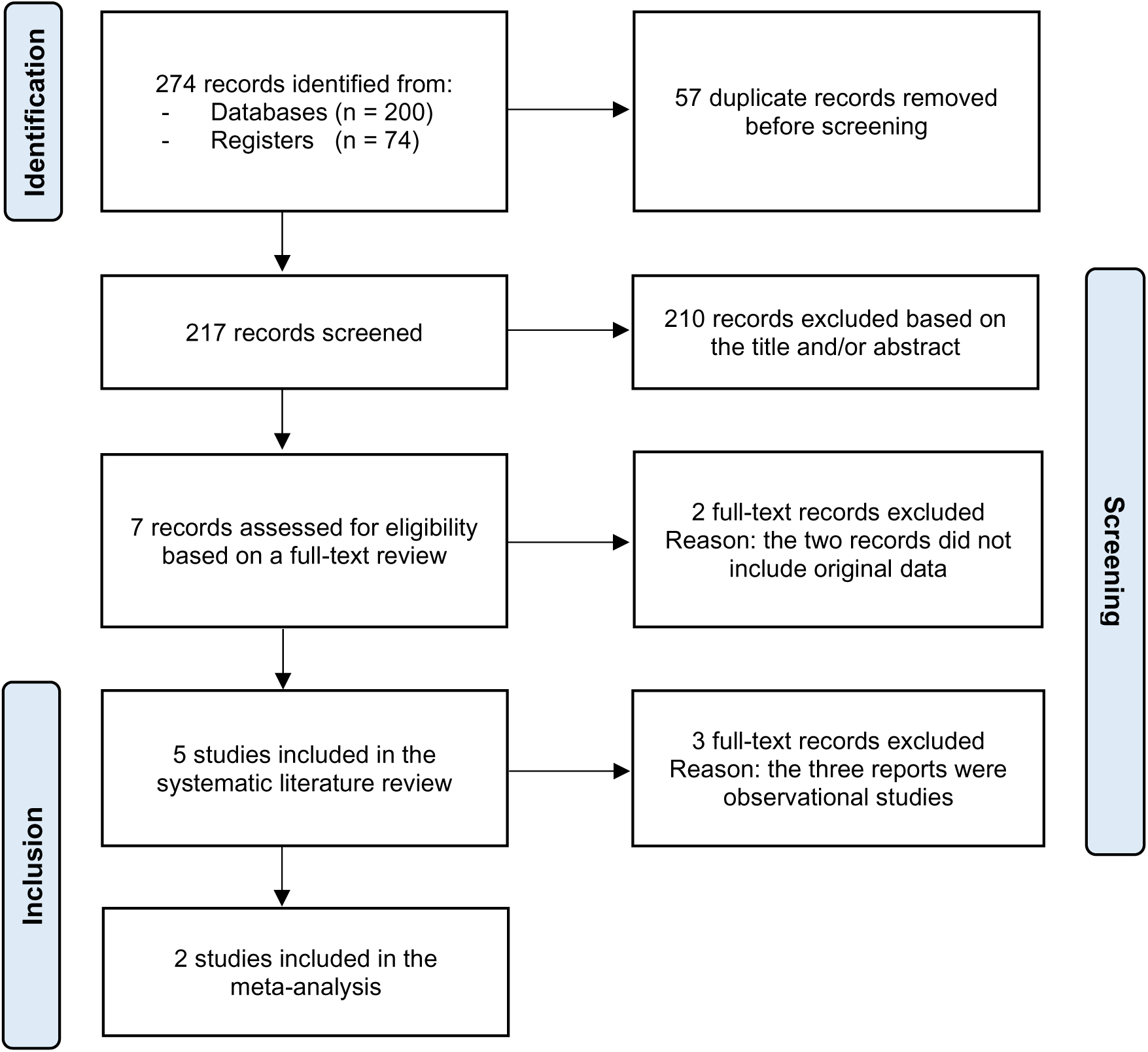
The PRISMA (Preferred Reporting Items for Systematic Reviews and Meta-Analyses) flow diagram illustrating the study selection process [25].

**Table 1:**
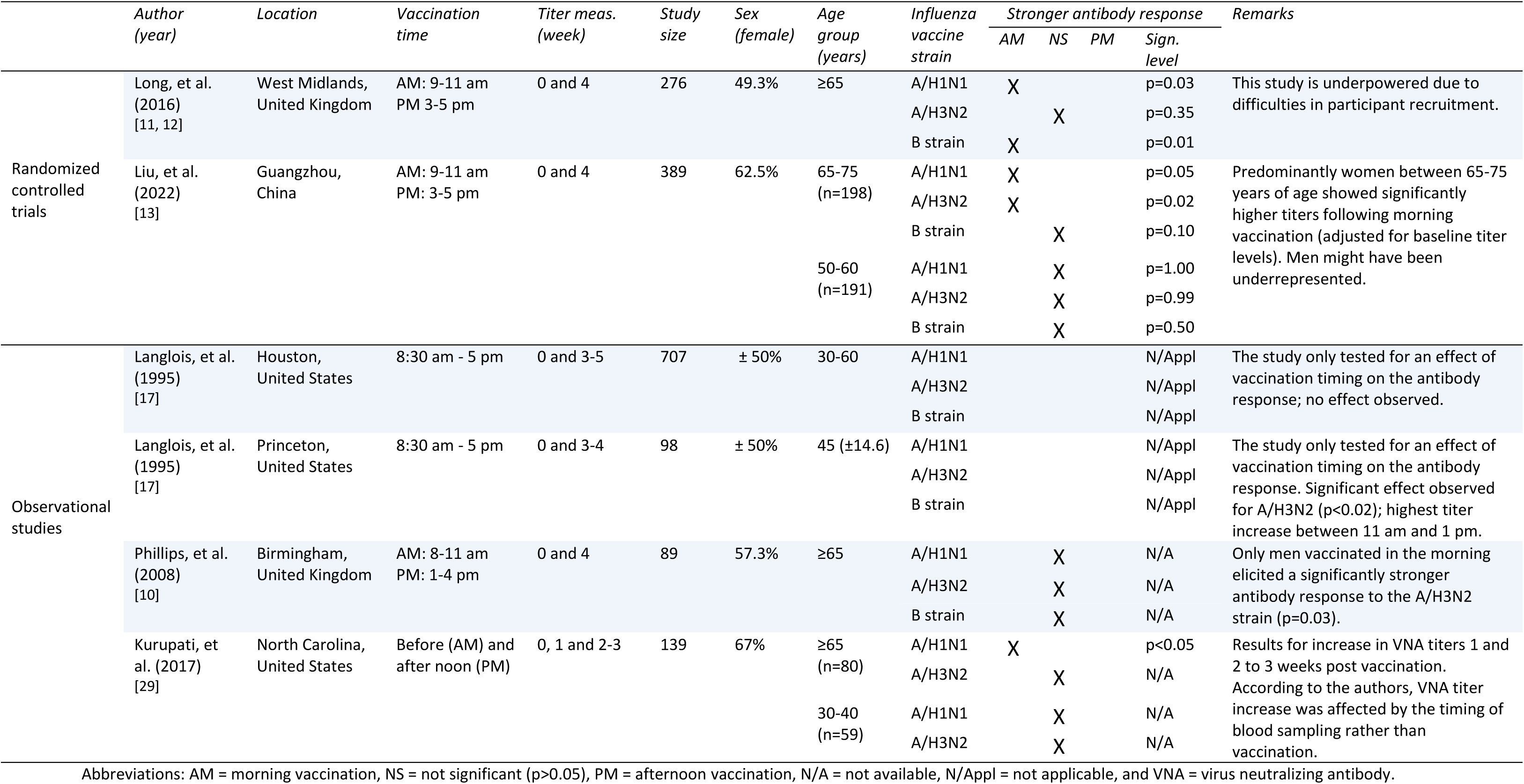
Characteristics and findings of the studies included in the systematic literature review.

### Systematic review

#### The timing of influenza vaccination impacts the antibody response

Two experimental studies by *Long, et al. (2016)* and *Liu, et al. (2022)* provided evidence for a causal relationship between vaccine administration timing and the strength of the antibody response. Both RCTs demonstrated that administering the influenza vaccine in the morning induces a stronger antibody response than afternoon vaccination in adults aged 65 years or older (Table 1) [11–13]. The study by *Long, et al. (2016)* was conducted over three influenza seasons, with slight antigenic variations in the three vaccine strains each year. Antibody responses to these strain variations were combined for the analysis, while the vaccine type was taken into account as a covariate. The inclusion of vaccine type did not influence the results, suggesting that this time-of-day effect extends to the annual variations in the influenza vaccine [11, 12]. *Liu, et al. (2022)* found that this time-of-day effect depended on sex. Postvaccination titers were significantly higher for women than for men aged 65-75 years (Table 1) [13].

An observational study by *Phillips*, *et al. (2008)* observed that men (aged ≥65 years) vaccinated in the morning had significantly higher anti-A/H3N2 antibody titers (Table 1) [10]. Women had a stronger antibody response following afternoon vaccination, but this lacked statistical significance [10]. These observed differences between sexes appear to contradict the outcomes of the two RCTs; *Long, et al. (2016)* found no differences between men and women, while *Liu, et al. (2022)* demonstrated that morning vaccination only resulted in a stronger antibody response in women [10].

*Kurupati, et al. (2017)* reported that older adults (aged ≥65 years) vaccinated in the morning had greater increases in antibody titers against the A/H1N1 and A/H3N2 vaccine strains than those vaccinated in the afternoon (Table 1) [29]. However, the authors attributed these differences to the timing of blood sample collection rather than the timing of vaccination [29]. The lack of standardization in the blood sampling time meant that those vaccinated in the morning were often also bled in the morning, and vice versa. Baseline antibody titers were consistently higher in the afternoon than in the morning, which resulted in a greater fold increase for individuals vaccinated in the morning compared to those vaccinated in the afternoon [29]. No differences in antibody titers were observed among adults aged 30-40 years based on the timing of vaccine administration or blood sample collection [29].

The study by *Langlois, et al. (1995)* analysed data from two influenza vaccination studies conducted in Houston and Princeton (USA) [17]. Participants of the Houston and Princeton studies were adults with mean ages of 43.9 (± 0.9) and 45.0 (± 14.6) years, respectively. Instead of categorizing participants into morning and afternoon vaccination groups, *Langlois, et al. (1995)* used multiple estimated time intervals and the actual (continuous) time of vaccine administration in their analyses for the Princeton and Houston study, respectively [17]. In the Houston study no association was observed between the antibody response and the time of day of vaccine administration [17]. The Princeton study observed significant variation in the 3-4 week increase in anti-A/H3N2 titers throughout the day. The greatest increase from baseline titers was observed for those vaccinated between 11 am and 1 pm, while those vaccinated at approximately 8:30 am and 5 pm had the lowest increase in anti-A/H3N2 antibody titers (Table 1 and Figure S3) [17]. After revaccination the following year, there was no longer a significant association between the anti-A/H3N2 response and the time of vaccination, suggesting that influenza vaccination history could mitigate this time-of-day effect [17].

#### Risk of bias in the included studies

The overall risk of bias was low for the RCTs, but varied across the observational studies (Figure S4). *Langlois, et al. (1995)* was the only study with a serious risk of bias, primarily due to the retrospective classification of the Princeton study participants into multiple vaccination time groups, which relied on the order of vaccination and the assumption that participants were continually vaccinated throughout the day. The included studies did not take the time of blood sample collection into account in their analyses, except for *Kurupati, et al. (2017)*. However, it remains unclear whether this could have affected the results.

### Meta-analysis

#### Estimating the overall effect of influenza vaccination timing on the antibody response

The antibody titers after morning and afternoon vaccine administration from the two RCTs were compared using the SMD, as illustrated in Figure 2. All effect sizes were positive, indicating that morning vaccination consistently resulted in higher antibody titers than afternoon vaccination. The pooled SMD was 0.24 (95% CI= 0.01–0.47, Z=2.07, p=0.038). A substantial level of heterogeneity was detected between the studies (*tau^2^*=0.023; *Q*=8.74; df=5; p = 0.12; *I^2^*=66%). Subgroup analyses revealed that the effect of vaccination timing was significantly stronger among adults aged ≥65 years (SMD=0.32, 95% CI: 0.21–0.43) than among those aged ≤60 years (SMD=0.00, 95% CI: −0.16–0.16, respectively). There were no statistically significant differences based on sex or between the influenza vaccine strains (Table 2).

**Figure 2:**
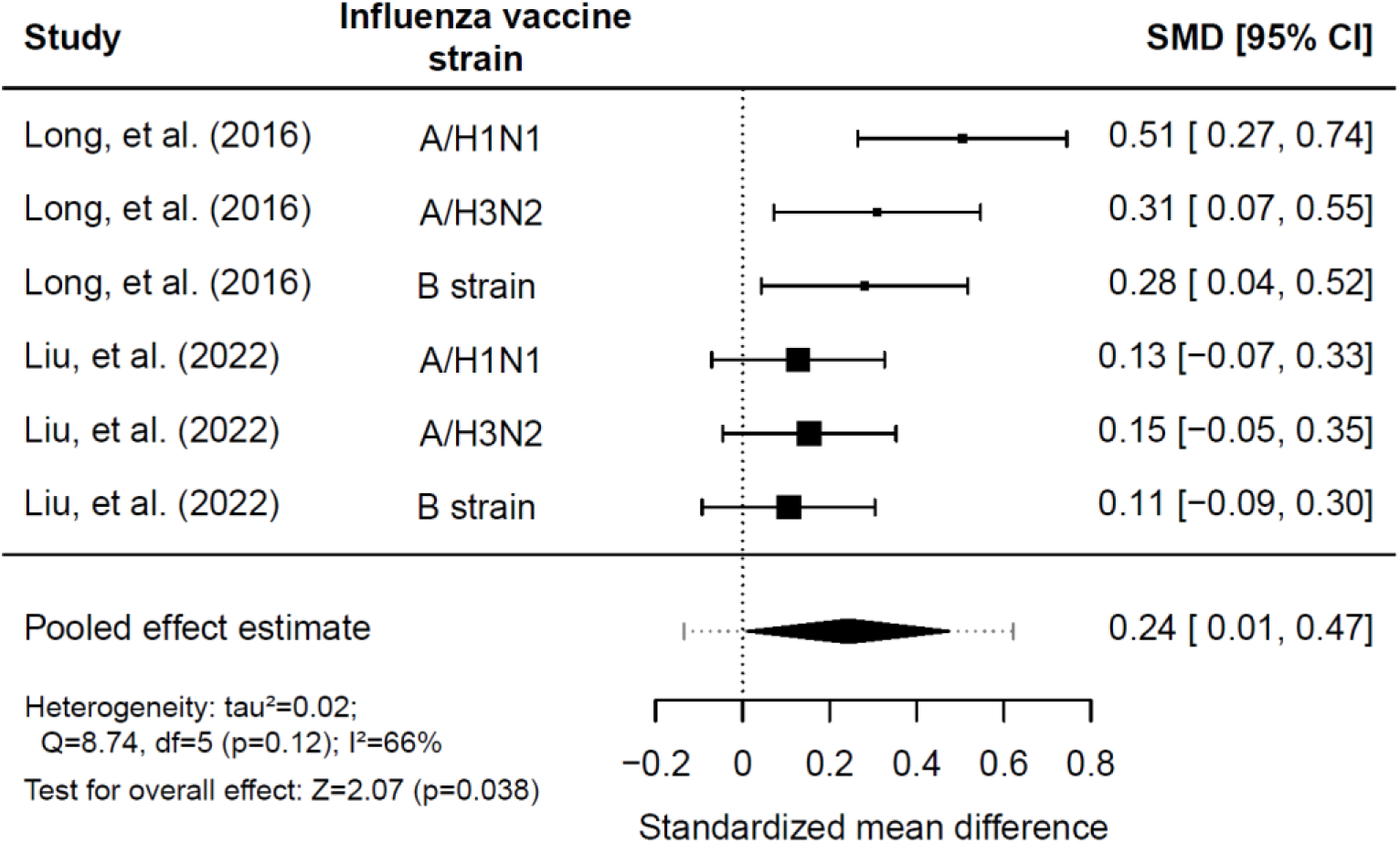
Comparison of log-antibody titers one month post vaccination between morning and afternoon administration of the influenza vaccine. SMD = standardized mean difference.

**Table 2:** Subgroup analysis for the comparison of log-antibody titers between morning and afternoon influenza vaccination.

| Subgroup | SMD (95% CI) | p-value |
| --- | --- | --- |
| Age: ≥65 years old | 0.32 (0.21 – 0.43) |  |
| Age: ≤60 years old | 0.00 (-0.17 – 0.17) | <0.0001 |
| Male | 0.17 (-0.01 – 0.42) |  |
| Female | 0.29 (0.04 – 0.54) | 0.1669 |
| A/H1N1 | 0.30 (0.04 – 0.56) |  |
| A/H3N2 | 0.24 (-0.03 – 0.50) | 0.5573* |
| B strain | 0.20 (-0.07 – 0.46) | 0.3421* |

\* Anti-A/H1N1 titer levels were used as the reference. SMD = standardized mean difference.

#### Publication bias

Clinical trial registers were checked for ongoing studies and studies that were completed but not published due to the lack of a significant effect of vaccination timing. No such studies were identified.

## Discussion

Morning administration of influenza vaccines consistently resulted in stronger antibody responses when a statistically significant effect of vaccination timing was observed. The time-of-day effect was only statistically significant for adults aged 65 years or older. According to the meta-analysis, this age group demonstrated higher strain-specific antibody titers one month following morning compared to afternoon vaccination, with a small-to-medium pooled SMD of 0.32 (95%CI: 0.21–0.43). No difference was observed in antibody responses among adults aged 60 years or younger (SMD=0.00). The RCTs included in the meta-analysis standardized the time of blood sample collection at the one-month follow-up visit [11–13], therefore we can infer that the difference in the postvaccination titers reported here is not attributable to variations in the timing of blood sampling. Randomization of the time of vaccination in these studies further substantiates the existence of a causal relationship between vaccination timing and the observed differences in antibody titers.

Our study raises the question why an improved morning vaccine response is observed only in adults aged 65 years or older. *Liu, et al. (2022)* speculated that this could be a result of immunosenescence, i.e. the gradual age-related decline in the function of both innate and adaptive immune responses [13, 30]. In younger adults the effect of vaccination timing may be concealed by their overall stronger immune response. The overall weaker immune response of older adults might be more sensitive to circadian oscillations in the immune system, which could result in age-dependent differences. While data for adults aged 61-64 years is unavailable, it is expected that the time-of-day effect varies gradually across the age groups and does not have a specific cut-off point at the age of 65 years. Furthermore, the studies reported conflicting results on sex-based differences for the effect of vaccination timing [10–13]. According to our meta-analysis, the effect of morning vaccination on the antibody response was larger for women than for men, albeit without statistical significance. Future research should further investigate differences in the effect of vaccination timing between men and women and explore sex-specific variations in circadian rhythms of the immune system.

A limitation of this study is the limited number of RCTs included in the meta-analysis. The paucity of relevant trials in this field underscores the need for additional research to better estimate the effect of optimizing vaccination timing for influenza and other infectious diseases. Our study suggests various ways in which future trials can improve upon the existing ones. Future trials should treat the time of vaccine administration as a continuous variable, as *Langlois, et al. (1995)* demonstrated that the optimal time for influenza vaccination might be between 11 am and 1 pm (Figure S3) [17]. The studies in the meta-analysis did not incorporate these timepoints within their morning and afternoon vaccination groups, which suggests that a larger effect of vaccination timing might be missed due to the comparison of two suboptimal time periods. Further research should also look into the effect of the timing of blood sample collection on antibody titer levels, which could lead to valuable insights for the design and analysis of serological studies. Furthermore, there is a lack of translation for the difference in the antibody response between morning and afternoon vaccination to a difference in the level of protection. To gain a better understanding of how vaccination timing affects the level of protection, further research should investigate the impact of vaccination timing on T-cell and long-term antibody responses.

Circadian oscillations in the immune system are a plausible cause of these time-dependent variations in the vaccine response [2, 9]. Therefore, the optimal time of day for vaccine administration might vary between individuals with different chronotypes (typically classified as morning, intermediate or evening types), as they exhibit inherent variations in the circadian phase of their biological clock [31]. Considering an individual’s circadian phase, rather than just the time of day, might provide a more accurate prediction of the optimal time for vaccine administration. However, the effect of chronotype on the optimal vaccination time has not received attention in current studies.

The generalizability of our findings to other vaccine types is a question for future research. Reports on the optimal timing for vaccines against other infectious diseases vary. Two other RCTs with young/middle-aged adults, conducted by *Karabay, et al. (2007)* and *Lai, et al. (2023)*, reported no significant differences in antibody titers between morning and afternoon vaccination against hepatitis B and SARS-CoV-2, respectively [18, 19]. Two observational studies [14, 15] and one partially randomized trial [10] reported more robust immune responses following morning administration of the following vaccines: inactivated SARS-CoV-2 [14], Bacillus Calmette-Guérin (BCG) [15], and hepatitis A [10]. The latter study found elevated antibody titers following morning vaccination against hepatitis A in young men, but not young women [10]. Furthermore, *Wang, et al. (2022)* demonstrated stronger anti-spike (SARS-CoV-2) responses in participants vaccinated later in the day with either mRNA or adenoviral vaccines [16]. This suggests that the optimal administration time may depend on the specific vaccine platform, potentially linked to the rate at which the vaccine antigens are recognized by the immune system.

### Implications and conclusion

Administering influenza vaccination in the morning, rather than the afternoon, induced a stronger antibody response in adults aged 65 years and older, potentially holding significant implications at the population level. This older demographic represents a major target group for influenza vaccination campaigns given their high rates of influenza-associated hospitalization and mortality [23]. Therefore, chrono-optimizing influenza vaccination could offer a simple strategy to boost vaccine effectiveness without incurring additional expenses or harm. Prioritizing morning vaccination for (most) adults of advanced age should be feasible and could be readily integrated into general practices. We hope that the evidence of the effect of influenza vaccination timing will spark more interest in investigating this time-of-day effect for vaccines against other infectious diseases. This could be facilitated if future vaccination trials record the timing of vaccine administration and take it into account when assessing vaccine efficacy.

## Availability of data and materials

Not applicable.

## Ethics approval, consent to participate and consent for publication

Not applicable.

## Authors’ contribution

JW and KV conceptualized and designed the study. JW supervised KV throughout the project. JK and KV conducted the literature search, screened the records, selected the articles and assessed the risk of bias in the included studies. JW was consulted if no consensus was reached for the study selection and risk of bias assessment. KV extracted the data and conducted the analyses. The manuscript was drafted by KV under the guidance of JW. All authors have read the manuscript and provided feedback. All authors reviewed the final version and granted approval for submission.

## Data Availability

All (aggregated) data used in this study were from the following articles:
Long, J.E., et al., Morning vaccination enhances antibody response over afternoon vaccination: A cluster-randomised trial. Vaccine, 2016. 34(24): p. 2679-85.
Liu, Y., et al., The impact of circadian rhythms on the immune response to influenza vaccination in middle-aged and older adults (IMPROVE): a randomised controlled trial. Immunity & Ageing, 2022. 19(1): p. 46.
Some data from Long et al. was collected via contacting the authors.

## Acknowledgements

We thank Jan van de Kassteele for the statistical advice regarding the meta-analysis.

## Abbreviations

RCT: Randomized controlled trial
PRISMA: Preferred reporting items for systematic reviews and meta-analyses
RoB2: Risk of bias tool 2
ROBINS-I: Risk of bias in nonrandomized studies of interventions
SD: Standard deviation
SMD: Standardized mean difference
BCG: Bacillus Calmette-Guérin

## Supplementary materials

**Table S1:**
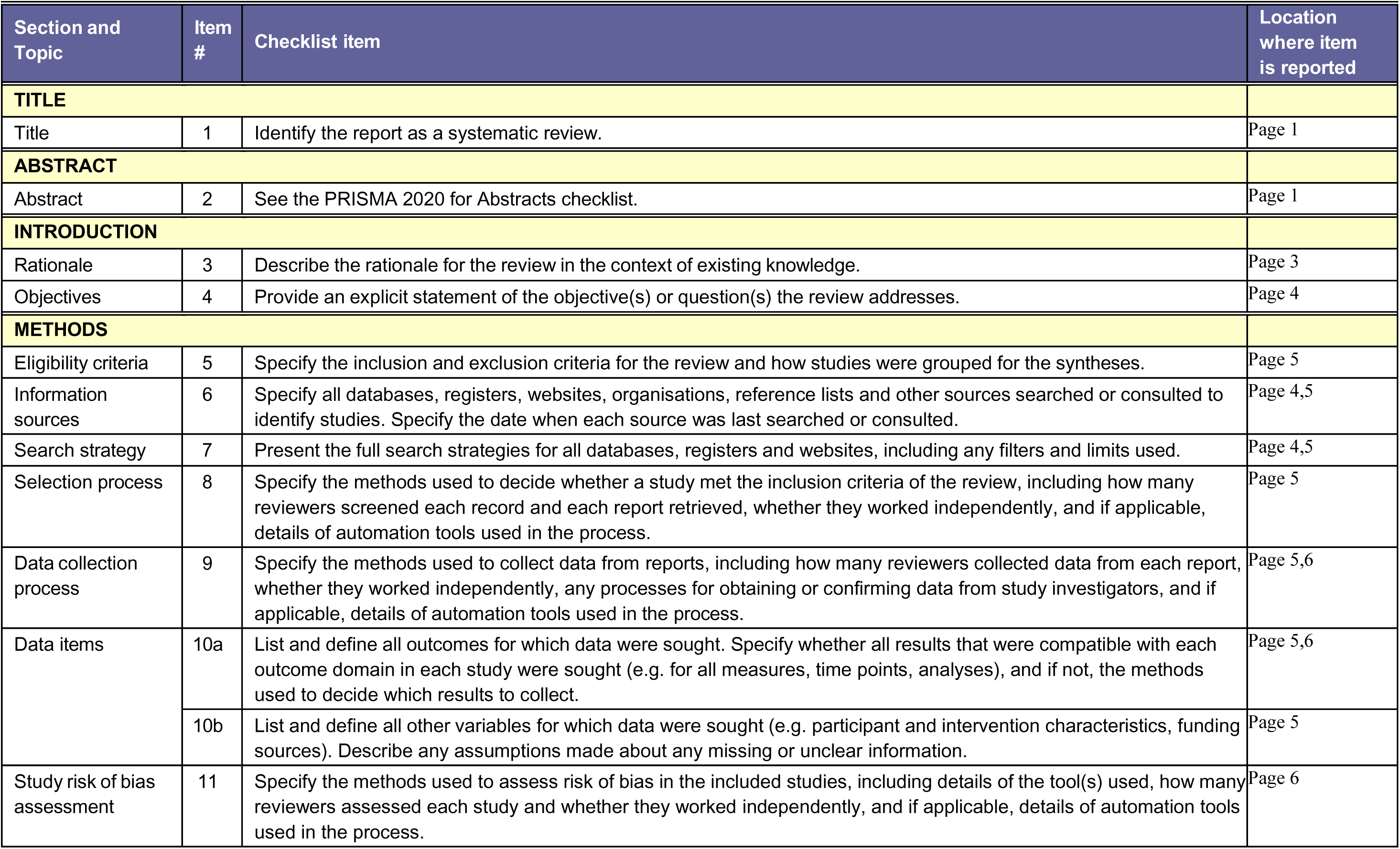
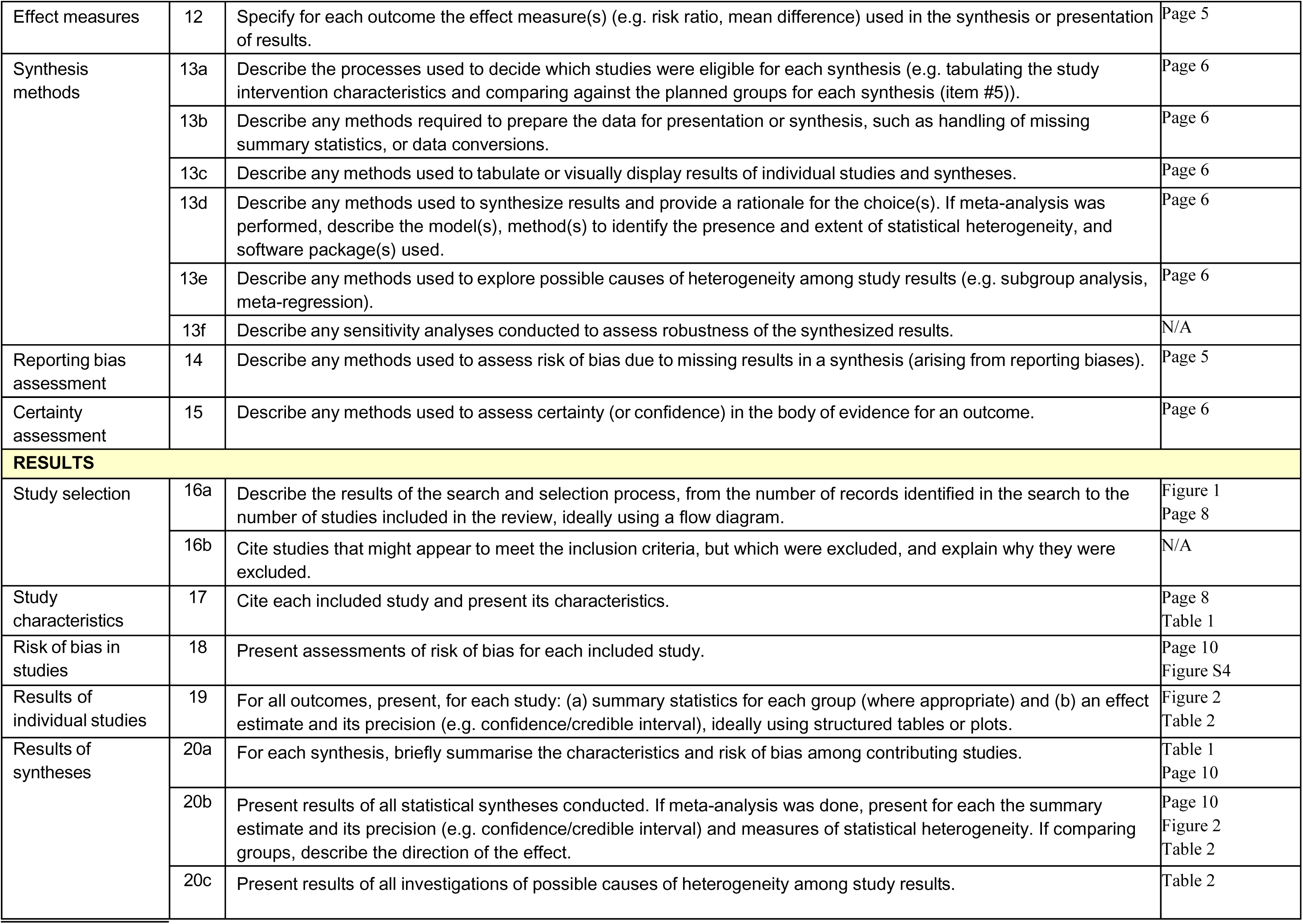
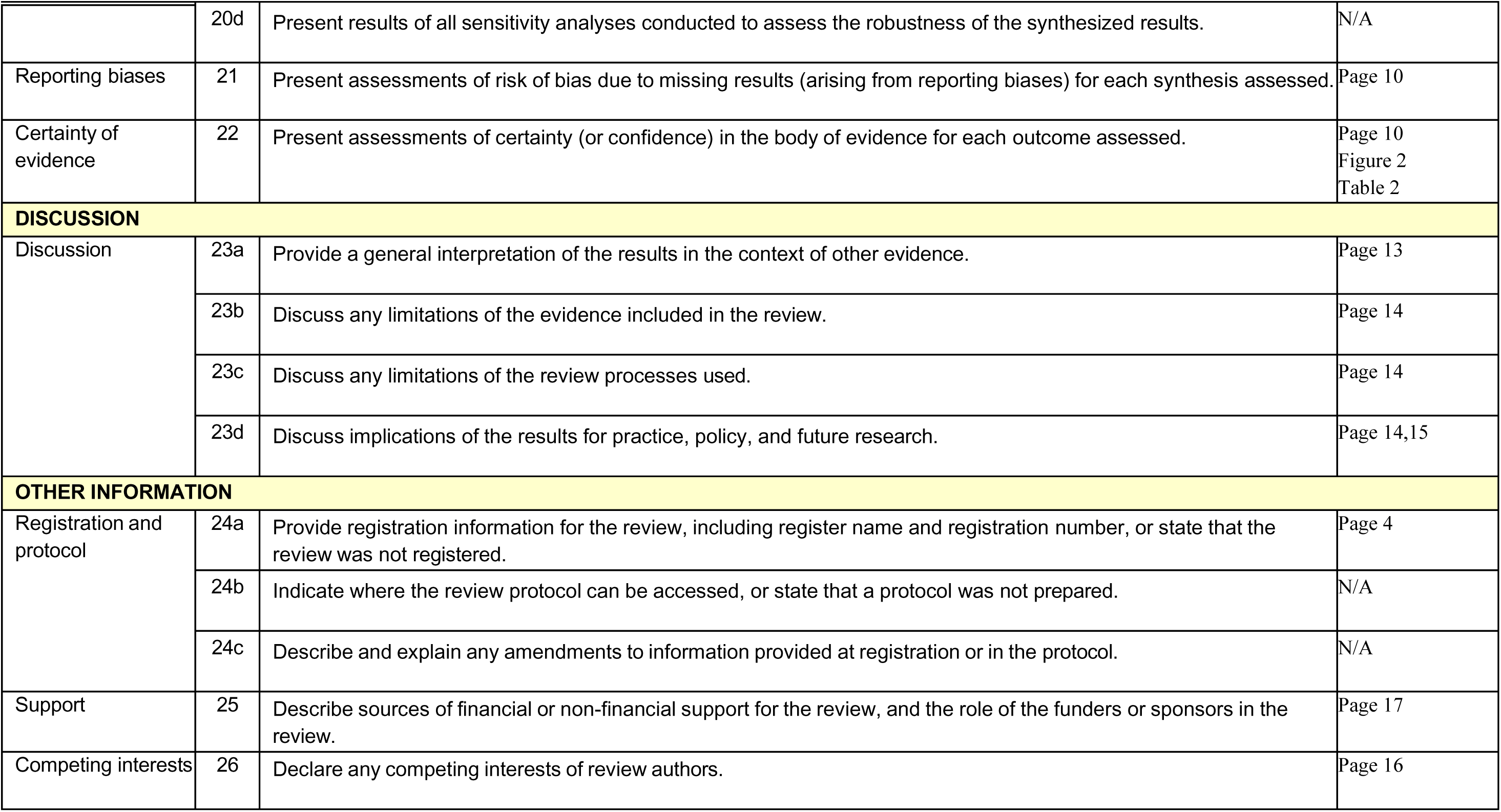
PRISMA 2020 item checklist [25].

### S2: Search string

The search strategy was based on the following search strings:

*Embase, MEDLINE and Preprints:*

((influenza AND vaccination OR (influenza AND vaccine)) AND vaccination AND timing OR (vaccination AND time) OR (time AND of AND day)) AND morning AND vaccination OR (afternoon AND vaccination)

*PubMed and Cochrane Central:*

((((((influenza vaccination) OR (influenza vaccine)) AND (vaccination timing)) OR (vaccination time)) OR (time of day)) AND (morning vaccination)) OR (afternoon vaccination)

Note that all word variations for the search terms have been used in the literature search. The full search string is available upon request.

**Figure S3:**
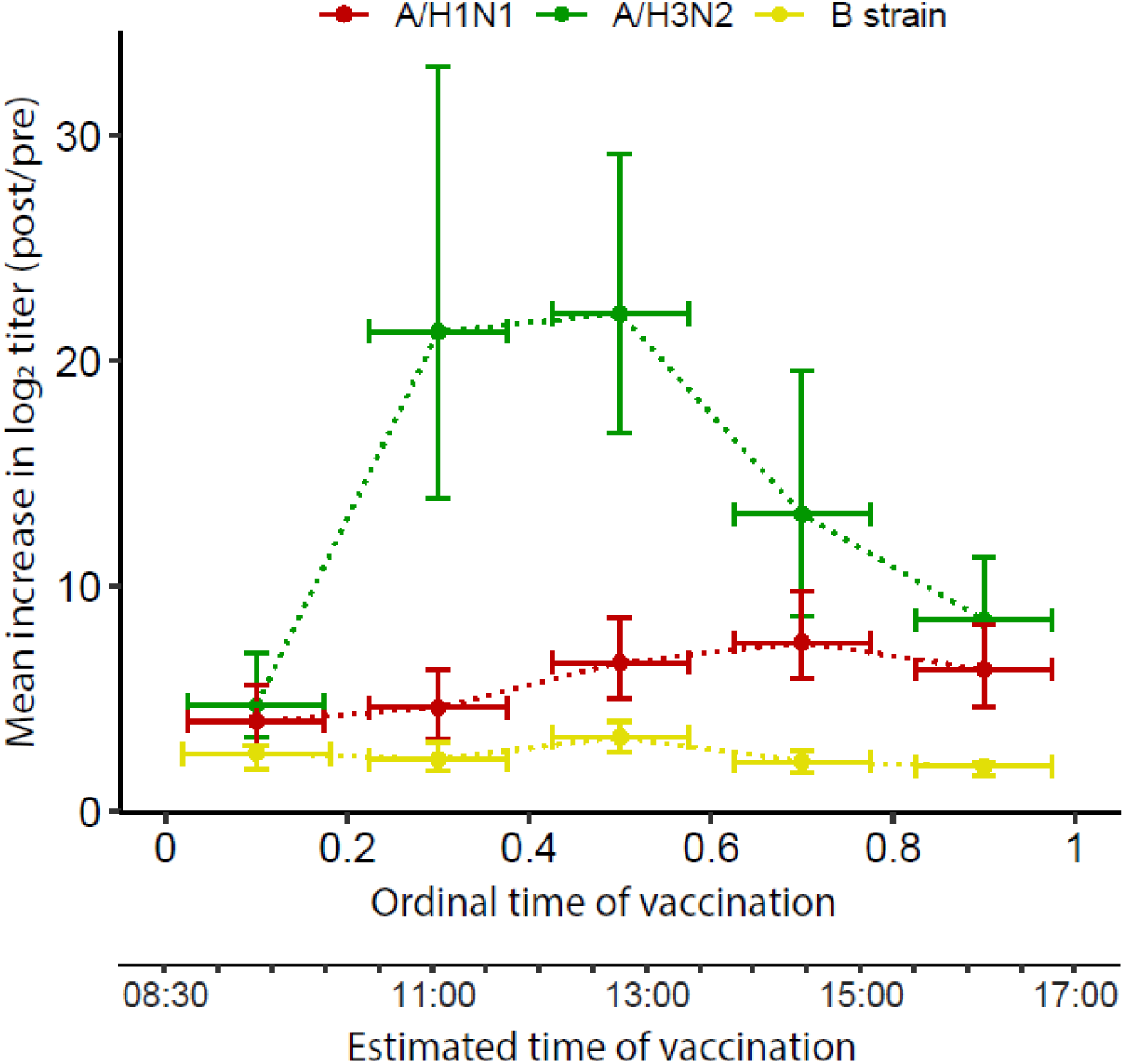
Redrawn figure of *Langlois, et al.* (1995) [17] illustrating the difference in the mean rise in log_2_ titer against the three influenza vaccine strains for multiple time groups. Vaccination time was estimated by the order in which the participants were vaccinated during clinic hours (08:30-17:00).

**Figure S4:**
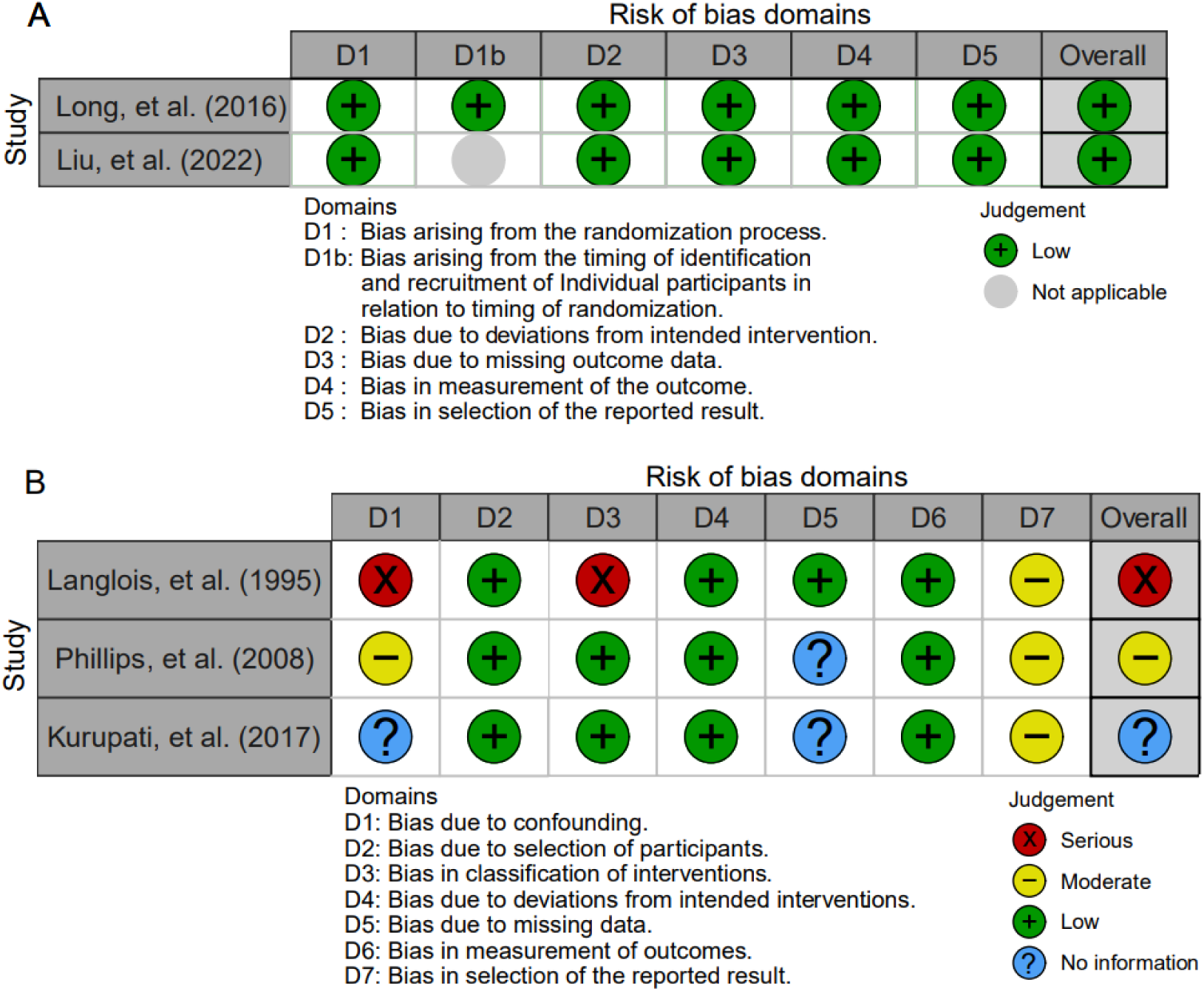
Risk of bias in the randomized clinical trials (**A**) and observational studies (**B**) included in the review and meta-analysis.

